# COVID-19 and maternal mental health: Are we getting the balance right?

**DOI:** 10.1101/2020.03.30.20047969

**Authors:** Anastasia Topalidou, Gill Thomson, Soo Downe

## Abstract

This paper presents a rapid evidence review into the clinical and psychological impacts of COVID-19 on perinatal women and their infants. Literature search revealed that there is very little formal evidence on the impact of COVID-19 on pregnant, labouring and postnatal women or their babies. The clinical evidence to date suggests that pregnant and childbearing women, and their babies are not at increased risk of either getting infected, or of having severe symptoms or consequences than the population as a whole. There is no evidence on the short- and longer-term psychological impacts of restrictive practices or social and personal constraints for childbearing women during COVID-19 in particular, or infection pandemics in general. The potential for adverse mental health consequences of the pandemic should be recognised as a critical public health concern, together with appropriate care and support to prevent and ameliorate any negative impacts.

## Introduction

Being pregnant and/or having a baby is, ideally, an event that is associated with joy, delight, and fulfilment, following a safe and positive pregnancy, birth, and early parenthood. However, some women and their partners can experience a range of negative emotions during this period, including anxiety and depression. Globally, the extent and adverse impacts of maternal mental health problems are increasingly recognised. As the World Health Organisation (WHO) states “*virtually all women can develop mental disorders during pregnancy and in the first year after delivery*”. Conditions such as extreme stress, emergency and conflict situations and natural disasters can increase risks for specific mental health disorders [1]. Maternal and parental mental health problems are associated with longer term risks for the mother and partner, and for their children [2] [3] [4] [5]. This raises the question: how are we safeguarding the short- and longer-term mental health of pregnant women and their partners in the age of coronavirus?

Apart from the overall population level pandemic-related stress, there is still a limited formal evidence-base about the nature and clinical consequences of the various versions of coronavirus (COVID-19 or SARS-CoV-2 or HCoV-19) for a pregnant woman. There is even less information about the mental health impacts consequent to self-isolation, living in a household with an affected person, limited access to goods/services and to routine or emergency health and social care. For women (and especially for those living in poverty, in poor or cramped housing), these issues may be exacerbated if they are expected to be the main carers for elderly or infirm relatives, or for young children, while living with multiple family members in confined spaces.

The most recent WHO guidelines for antenatal and for intrapartum care reinforce the importance of positive clinical and psychosocial pregnancy and childbirth experiences to optimise the physical and psychosocial wellbeing of mother, baby, and the family in the short and longer term [6] [7]. It is therefore critical that knowledge about the clinical impact of COVID-19 in pregnancy is balanced with knowledge about the potential psycho-social impact, so that the current rapid restructuring of maternity services to cope with the pandemic can take account of all the relevant consequences.

### What is currently known about COVID-19 and pregnancy?

A rapid scoping review of OVID in Embase [1974 to 2020 March 21], Ovid MEDLINE(R) and Epub Ahead of Print, In-Process & Other Non-Indexed Citations and Daily [1946 to March 21, 2020], Maternity & Infant Care Database (MIDIRS) [1971 to February 2020] databases generated 666 hits. This body of knowledge is based on case reports and observational studies, with small cohorts and consequent limitations [8] [9] [10] [11] [12] [13] [14] [15]. The key findings from these studies have been captured in a number of rapid COVID-19 guidelines produced by professional and global authorities. In summary, they are as follows:

#### Clinical findings

1. There is no evidence that the virus can be transmitted vertically from the mother to the fetus during pregnancy. This is based on; one study of 9 pregnant women with laboratory-confirmed COVID-19 pneumonia [8];a single case report [9]; a study of 41 pregnant women with confirmed COVID-19 [11]; and a study of 15 pregnant women with COVID-19 [10].
2. There is no reported transmission during birth. Infants born vaginally or by caesarean section, to mothers with coronavirus infection, have so far been shown to be negative to COVID-19. This is based on: an observational study of 4 mothers infected with COVID-19 and 3 newborns [12]; a single case report [9]; and an observational study on 11 mothers infected with COVID-19 and their newborns [10]. No virus was detected in the amniotic fluid, cord blood, neonatal throat swab and breastmilk samples (results from 6 patients with confirmed COVID-19) [8]. A recently reported single case of a neonate that tested positive for COVID-19 does not provide definitive evidence of intrauterine or intrapartum infection, as no intrauterine tissue sample was tested, and the neonate was tested 30 hours after birth. However, this case does suggest that cross infection can occur at some point between pregnancy and the early postnatal period [8] [13].
3. Despite this single case, the risk/benefit calculation is that mothers and babies should not be routinely separated after birth to prevent transmission of COVID-19. In the two published studies from China referenced above (total of 5 participants), infants were isolated from their mothers immediately after birth for up to 14 days [9] [12]. However, the guidelines from WHO for mothers infected with COVID-19 are “*Close contact and early, exclusive breastfeeding helps a baby to thrive*”, “*Hold your newborn skin-to-skin*” and “*share a room with your baby*” [16]. In addition, UNICEF recommendations during the period of the outbreak are “*Regardless of feeding method, it is essential that sick and preterm babies’ profound need for emotional attachment with their parents/primary caregiver continues to be considered. Keeping mothers and babies together wherever possible and responding to the baby’s need for love and comfort will not only enable breastmilk/breastfeeding, but will also protect the baby’s short- and long-term health, wellbeing and development. In addition, this will support the mother’s mental wellbeing in the postnatal period”* [17].
4. There is no evidence or biological indication that COVID-19 can be transmitted through breastmilk. As the benefits of breastfeeding are well-known, particularly in terms of boosting the infant’s immune system, and maternal and child bonding, WHO recommends that “*women with COVID-19 can breastfeed of they wish to do so”*. Given the potential for postnatal transmission indicated by the single case of neonatal infection noted above, WHO adds that women who are breastfeeding, and, indeed, those who are giving babies formula milk, should “*practice respiratory hygiene during feeding, wearing a mask where available, wash hands before and after touching the baby and routinely clean and disinfect surfaces they have touched*” [16].
5. In one large observational studies of 2143 paediatric patients with COVID-19 in China, one child (aged 14) died. Although the overall incidence of severe disease was highest in the under one year old group (10.6%) this was still much lower than in adult patients (18.5%) suggesting that babies and children are much less at risk of adverse effects of being infected with COVID-19 and much less likely to die as a result than adults. [14].
6. There is no evidence to date that pregnant women are more likely to acquire COVID-19 than other young adults, or that, once they have it they have more severe disease, or that those with mild or severe disease have a higher risk of fetal distress or of adverse fetal outcomes. This information is based on two small observational studies from China [10] [15].
7. Based on Royal College of Obstetricians and Gynaecologists there is no evidence “*that epidural or spinal analgesia or anaesthesia is contraindicated in the presence of coronaviruses. Epidural analgesia is therefore recommended before, or early in labour, to women with suspected/confirmed Covid-19 in order to minimise the need for general anaesthesia if urgent delivery is needed*” [9] [18].

The above information may change as knowledge evolves on a daily basis.

#### Psychological findings

No studies were found for COVID-19 relating to maternal mental health, and to the best of our knowledge there are no studies or case reports that explored the influence of other epidemics or pandemics such as H1N1 influenza or 2014-16 Ebola on the mental health of expectant, or new mothers.

### No health without mental health

This gap is critical. It is especially important when two of the Millennium Development Goals (4 and 5) emphasise on maternal and child health, stating that overall health cannot be achieved without mental health [2]. Psychosocial stress, particularly during early pregnancy, has been identified as a key risk factor in the aetiology of a preterm birth [19]. The potential (ill-informed) practices of separating mothers from their infants post birth in the name of protection from COVID-19 infection, such as witnessed in China, and a lack of certainty leading to early cessation of breastfeeding, have harmful impacts on maternal mental health via ‘toxic stress’ [20] and perceptions of self-blame and shame [21]. The links between social isolation and depression are well reported [22], and potentially magnified when coupled with juggling the demands of confined family members. A recent article titled *The Coronavirus Is a Disaster for Feminism* also highlights how women’s independence is the ‘silent victim’ of the pandemic due to women being more likely than males to take responsibility for caregiving activities [23]. This situation is even more acute for women who face complex life issues such as poverty, or domestic violence. Women may be exposed to an increased risk of domestic violence, or who are trying to protect children from risks of abuse from relatives who are now confined at home; with a 300% increase in domestic violence cases reported in China during the pandemic [24]. Recent headlines also report further impacts of the COVID-19 crisis on women’s choices, such as Texas banning abortions as a non-essential operation – with little consideration of the mental fallout and wider implications of this decision-making [25].

Even though there is plenty of scientific evidences and experience-based knowledge in several fields from the previous two coronaviruses, the research on MERS-CoV and SARS-CoV and pregnancy/childbirth is still limited, especially in terms of maternal mental health. There is a critically important gap in our knowledge about how pandemics affect mothers and their babies, and how pregnant women, mothers and their families can be better supported. In February 2020, several reports were published in The Lancet stating that mental health care should be included in the national public health emergency systems and that further understanding was needed to better respond to future unexpected disease outbreaks [26] [27] [28] [29] [30]. Prenatal and postnatal mental health should be prioritised due to its pervasive short and long-term impacts on maternal, familial and fetus, infant and child biopsychosocial development [31].

## Conclusion

Current evidence suggests that pregnant women and babies are no more at risk of catching COVID-19 than other members of the public, and that there is no evidence that pregnant women are more likely to experience severe illness than other young adults, or that even severe illness is more likely to be associated with adverse neonatal outcomes. While there are a very few cases in which maternal to infant transmission may have occurred in utero this evidence is not definitive. Policies that are put in place to restrict maternal choices and rights in the name of protection of mother and/or baby are therefore not justified based on current evidence, and are likely to be associated with longer term harms, both physical and psychological. Services should not enforce unnecessary practices in the name of protection, including separation of mother and baby, or of chosen pain relief methods, and recommendations not to breastfeed should be actively avoided.

The lack of knowledge about the short- and longer-term psychological wellbeing of mother and baby following their experiences of maternity care during a pandemic is a serious gap in knowledge for optimal maternity care at this time. In the absence of such formal evidence, the potential for adverse mental health consequences of the pandemic should be recognised as a critical public health concern, together with appropriate care and support to prevent and ameliorate any negative impacts.

## Data Availability

Not applicable. This article is a commentary.

